# Genome surveillance of SARS-CoV-2 variants and their role in pathogenesis focusing on second wave of COVID-19 in India

**DOI:** 10.1101/2022.01.28.22269987

**Authors:** Poulomi Sarkar, Sarthak Banerjee, Sarbar Ali Saha, Pralay Mitra, Siddik Sarkar

## Abstract

India had witnessed unprecedented surge in SARS-CoV-2 infections and the dire consequences during the second wave of COVID-19, but the detailed report of the epidemiological based spatiotemporal incidences of the disease is missing. Here in, we have applied various statistical methods like correlation, hierarchical clustering to know the pattern of pathogenesis of the circulating VoCs. B.1.617.1 (Kappa) was the predominant VoC during the early phase of second wave. Delta (B.1.617.2) or Delta-like (AY.x) VoC constitutes majority (>90.17) of the cases during the peak of second wave. The correlation plot showed Delta/Delta-like lineage is inversely correlated with other lineages including B.1.617.1 (kappa), B.1.1.7, B.1, B.1.36.29 and B.1.36. Delta/Delta-like surge coincided with second wave whereas all other lineages (B.1.617.1, B.1.36.29, etc.) occurred during the prior phase of the second wave. The spatiotemporal analysis showed that most of the Indian states were affected during the peak of the second wave due to delta surge and fall under the same cluster. The second cluster populated mostly by north-eastern states and islands of India were minimally affected. The presence of signature mutations (T478K, D950N, E156G) along with L452K, D614G and P681R within the spike protein of Delta or Delta-like might cause elevation in host cell attachment, increased transmission and altered antigenicity which in due course of time has replaced the other circulating variants. The timely assessment of new VoCs will provide a rationale for updating the diagnostic, vaccine development by medical industries and decision making by various agencies including government, educational institutions, and corporate industries.

## Introduction

COVID-19 is the largest ongoing public health emergency since the 1918 influenza pandemic (1) which has claimed over 5.2 million of lives while infected >260 million population globally (2). The first COVID-19 case in India was reported in January 2020 which quickly spread all over the country and as of November 2021 the total infected population is >34.4 million, while >400 thousand people have succumbed to the disease (3). Lack of precise antiviral drugs is a major challenge in restricting the virus from disease spread. The development and increased use of safe vaccines may prove promising in future eradication of SARS-CoV-2 (2). But due to continuous mutation of RNA viruses including SARS-CoV-2, the efficacy of vaccines against the newly derived lineages than that of reference genome (against which the vaccines were designed) might be the concern (4) in near future. Genomic and epidemiological surveillance have been seen as a gold standard for control of contagious diseases. It helps in containing the transmission of the virus by identifying different novel viral variants (2). SARS-CoV-2 acquire genetic mutations similar to other RNA viruses (2) and these alterations lead to genesis of more communicable variants or variants of concern (VoC). Genome analysis of SARS-CoV-2 revealed that the Spike protein including Receptor Binding Domain (RBD) of homo-trimeric spike glycoprotein are altered in these communicable VoC, responsible for the spread and prolong COVID-19. The RBD of Spike protein participates in attachment to host cell ACE-2 receptor, thereby triggering an array of reactions for viral entry into the host cell (5). Mutations in the RBD region are found to be responsible for increased activity of ACE-2 (6), subsequently leading to a massive surge in the infection rates. Here we aimed at understanding the evolution of different variants of SARS-CoV-2 in different parts of India during the onset and subsidence of second wave (January-September, 2021) using genome based data. Owing to the sudden emergence of B.1.1.529/ BA.* (Omicron-like; first detected in Hong Kong/ South Africa) variant, and few cases registered in India, it was also included in our study. The study highlights about different lineages and VoCs circulating in Indian masses, their dynamic distribution in various Indian states during the onset, peak and post second wave in India.

## Materials and Methods

### Data mining and plotting

The SARS-CoV-2 genome sequences and patient metadata (n=44514) were retrieved from GISAID SARS-CoV-2 database. The nucleotide FASTA and patient metadata files were downloaded and used for the different analyses as mentioned in Supplementary Section.

### Spatiotemporal transmission/incidences of Delta in India

The spatial polygonal regions of different states and union territories (UTs) were obtained as described in detailed in supplementary methods. The shape file of the state level in India was obtained and downloaded from the [GISMAP](https://map.igismap.com/gis-data/india/administrative_boundaries_state_level). The metadata file obtained from GISAID databases for B.1.617.2 (Delta) incidences per month is analysed with respect to different states and UTs and plotted using R/ R studio with ggplot2 and relevant packages. The R code can be shared on request.

## Results

### COVID-19 cases and associated deaths across the world in 2021

There are 279114972 (279 million) cumulative COVID-19 cases across the world population and death associated with COVID-19 are 5397580 accounting on average 19338 (1.93 %) deaths per million COVID-19 cases. In India, there are 13797 (1.34 %) deaths reported per million COVID-19 cases and there are 15643 (1.56 %) deaths per million reported cases in USA. The detailed total cumulative cases, deaths or deaths per million reported cases are shown in Figure 1A, B and C, and Supplementary Table S1). Based on cumulative cases as of December 2021, USA, India and Brazil have reported the highest number of cases both in terms of cumulative cases and cumulative deaths. Interestingly, the deaths per million reported cases are found to be more in low- income and middle low income countries like Yemen, Sudan, Peru, Mexico, Syria, etc. Box plot clearly indicates that cumulative COVID-19 cases are more in developed nation with high-income group. It might be due to developed infrastructure and testing centers for COVID-19. Although the reported cases are lower in low-income countries, but deaths associated with COVID-19 are relatively higher in poor nations (Figure 1D-E). It might be due to lesser testing centers, poor medical facilities and social awareness. India, the country with estimated population of 1.3 × 10^9^, and falling under category 4; lower-middle income is expected to be affected by pandemic COVID-19 as shown in Figure 1. Here in, we focused on India, which has faced the catastrophic consequences of the second wave (April-June 2021) as shown in Supplementary Figure S1with peak during the month of May 2021. The distribution of cases and deaths related to COVID-19 in India in 2021 also depicts the peak during May (Supplementary Figure S1).

**Fig. 1.**
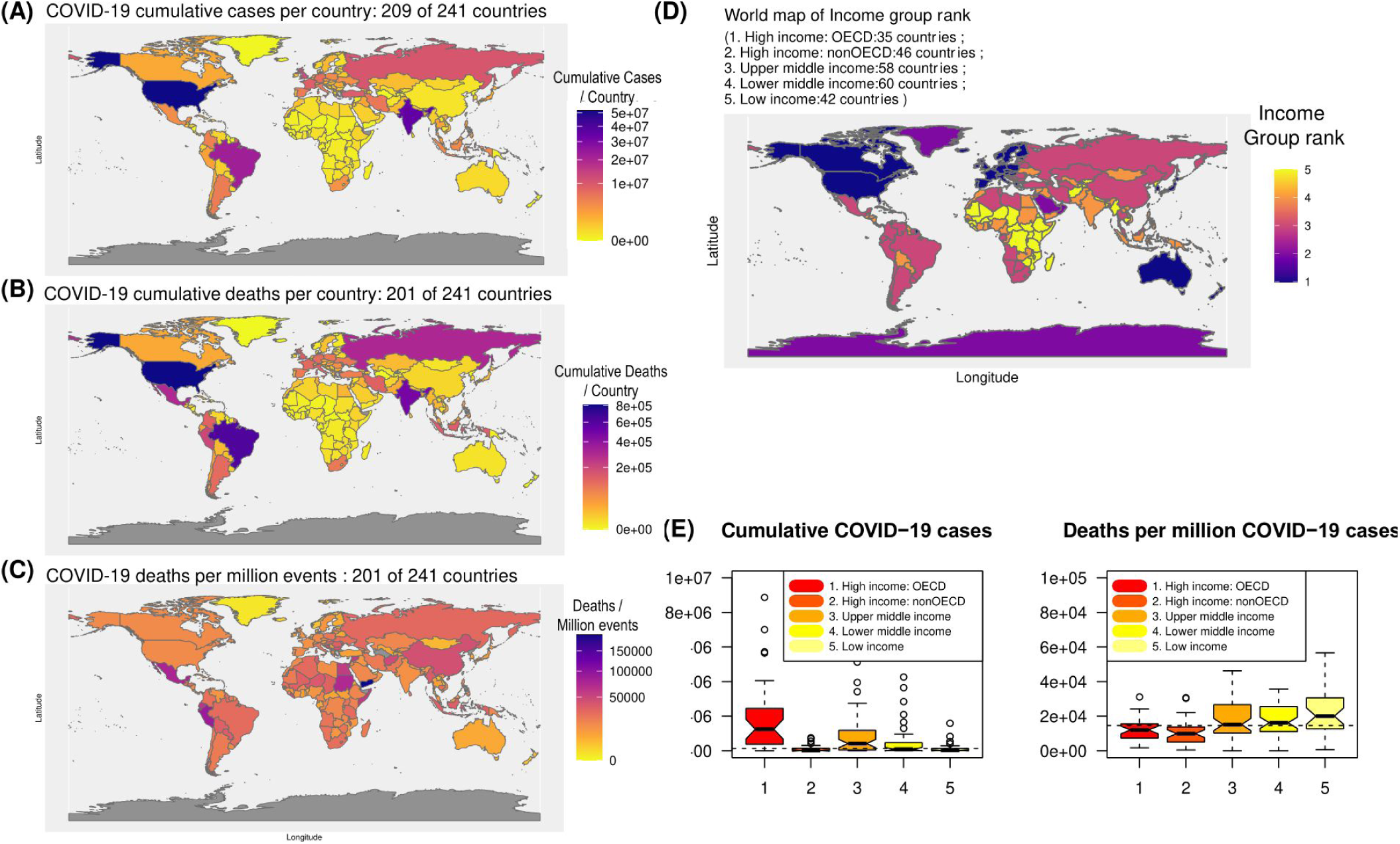
COVID-19 in world scenario in 2021. *A*; Cumulative cases of COVID-19 per country, *B*; Cumulative COVID-19 associated deaths per country, *C*; Cumulative deaths per million events (COVID-19) per country and *D*; Income group rank of the country is plotted in world map. *E* ; Box plot of COVID-19 cumulative cases or deaths with respect to income group ranking of the countries. *Note: The boundaries or geographical coordinates are used only for data representation. The administrative/geographical boundaries might differ*.

### Genome surveillance of SARS-CoV-2 in 2021 in India

To study the details and the possible variants of concern (VoC) of SARS-CoV-2 prevalent in India during the catastrophic 2nd wave, we retrieved patient metadata and the associated genome sequences (44797) from GISAID for the month of January (4 months prior to 2nd wave) to September, 2021 (4 months post 2nd wave). The respective metadata file (Supplementary Table S2) of all ‘Variants of Concern’ (VoC) and ‘Variants of Interest’ (VoI) as described in Methodology section were selected for the analysis. There are >=38 SARS-CoV-2 lineages found across India from January to September 2021 with minimum 100 recorded cases (Supplementary Table S3). The lineages were ordered in descending manner and top ten different variants of the virus as identified in India till September 2021 were plotted (Figure 2). Due to various sub-lineages of B.1.617.2 (Delta), AY.* (Delta-like) subseries was continuously renamed by Pango nomenclatures in GI-SAID. AY.4 got reclassified as AY.4.2, AY.12, AY.22, AY.23, AY.103, AY.122, etc. Hence, a generalized term AY.x is being used for the newly classified AY.4 variant. The graph was constructed based on percentage of incidences per month. The top 10 lineages found based on cumulative cases along with their prevalence (in %) per month was shown in Table 1 and Figure 2. Analysis of the plot depicts variants namely B.1, B.1.1, B.1.17, B.1.36 and B.1.36.29 were prevalent in the month of January. Likewise February showed similar pattern of occurrences except there was a rise in B.1. In March decline of all the other lineages could be seen while rise in the B.1.17, B.1.617.1 and B.1.617.2 (aka Delta variant) were reported. B.1, B.1.1.7, and B.1.617.1 remained prevalent prior to 2nd wave in India (Figure 2, Supplementary Figure S2). B.1617.2 surge/peak is found in the month May and coincides with the time of the second wave in India. B.1.617.2 in due course of time is further mutated to give rise to sub lineage of Delta-like AY.x. Both Delta/Delta-like constitutes >90 % of load of all cases per month during the peak and thereafter. During the post second wave, all other lineages including kappa which is prevalent during the onset or prior to second wave slowly diminishes and unable to compete with Delta/Delta-like lineages.

**Fig. 2.**
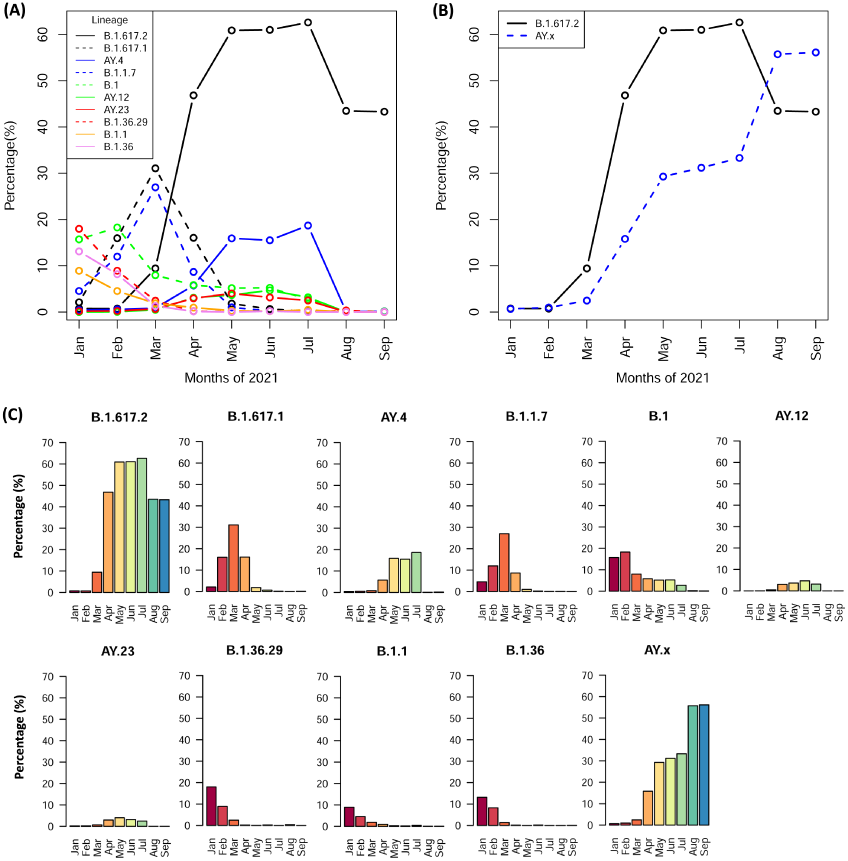
Genome Surveillance of top 10 VoC in India during Jan-Sept 2021. *A-B*; Scatter plot of incidences of various lineages as indicated per month. *C*; Bar plot of respective lineages as indicated.

**Table 1.** Incidences of reported cases of various VoC per month

| Month | B.1.617.2 | B.1.617.1 | AY.4 | B.1.1.7 | B.1 | AY.12 | AY.23 | B.1.36.29 | B.1.1 | B.1.36 | AY.x | B.1.617.2/AY.x |
| --- | --- | --- | --- | --- | --- | --- | --- | --- | --- | --- | --- | --- |
| Jan | 0.76 | 2.1 | 0.42 | 4.54 | 15.71 | 0 | 0.17 | 17.98 | 8.91 | 13.11 | 0.67 | 1.43 |
| Feb | 0.74 | 15.95 | 0.55 | 11.97 | 18.28 | 0.04 | 0.22 | 8.92 | 4.53 | 8.18 | 0.96 | 1.69 |
| Mar | 9.43 | 31.07 | 0.84 | 26.96 | 7.96 | 0.47 | 0.67 | 2.44 | 1.88 | 1.31 | 2.47 | 11.9 |
| Apr | 46.85 | 16.04 | 5.74 | 8.67 | 5.83 | 3.09 | 2.95 | 0.18 | 0.96 | 0.12 | 15.81 | 62.66 |
| May | 60.89 | 1.82 | 15.92 | 0.99 | 5.17 | 3.66 | 4.02 | 0.04 | 0.29 | 0 | 29.28 | 90.17 |
| Jun | 61.01 | 0.64 | 15.52 | 0.22 | 5.23 | 4.69 | 3.15 | 0.2 | 0.16 | 0.18 | 31.19 | 92.2 |
| Jul | 62.62 | 0.14 | 18.71 | 0.06 | 2.75 | 3.18 | 2.49 | 0 | 0.46 | 0 | 33.31 | 95.93 |
| Aug | 43.49 | 0.03 | 0.06 | 0.06 | 0.22 | 0 | 0 | 0.35 | 0.03 | 0 | 55.74 | 99.23 |
| Sep | 43.31 | 0.09 | 0.15 | 0.03 | 0.12 | 0 | 0 | 0 | 0.03 | 0.03 | 56.13 | 99.44 |

### B.1.617.2 (Delta) and AY.x (Delta like) distribution in Indian states and territories

In order to study the kinetics or virus spread which is mainly contributed by Delta lineage dynamic distribution of B.1.617.2 in different states during January to September, 2021 is plotted in Figure 3. The map showed that in January 5 states reported B.1.617.2 cases which increased to 29-32 during the peak of the second wave based on the reported cases in GISAID. From July onwards northern and central states of India reported declining B.1.617.2 cases while the case load borne by B.1.617.2 remained almost same in the southern part till September, 2021. Along with the Delta variant hike of AY.4/AY.x (aka Delta plus) was also eminent. AY.4/AY.x followed similar pattern as B.1.617.2 but with lesser incidences (% cases=13.98±4.01) in peak periods (Figure 2, Table 1, Supplementary Figure S3), but reached >50% of incidences per month by September.

**Fig. 3.**
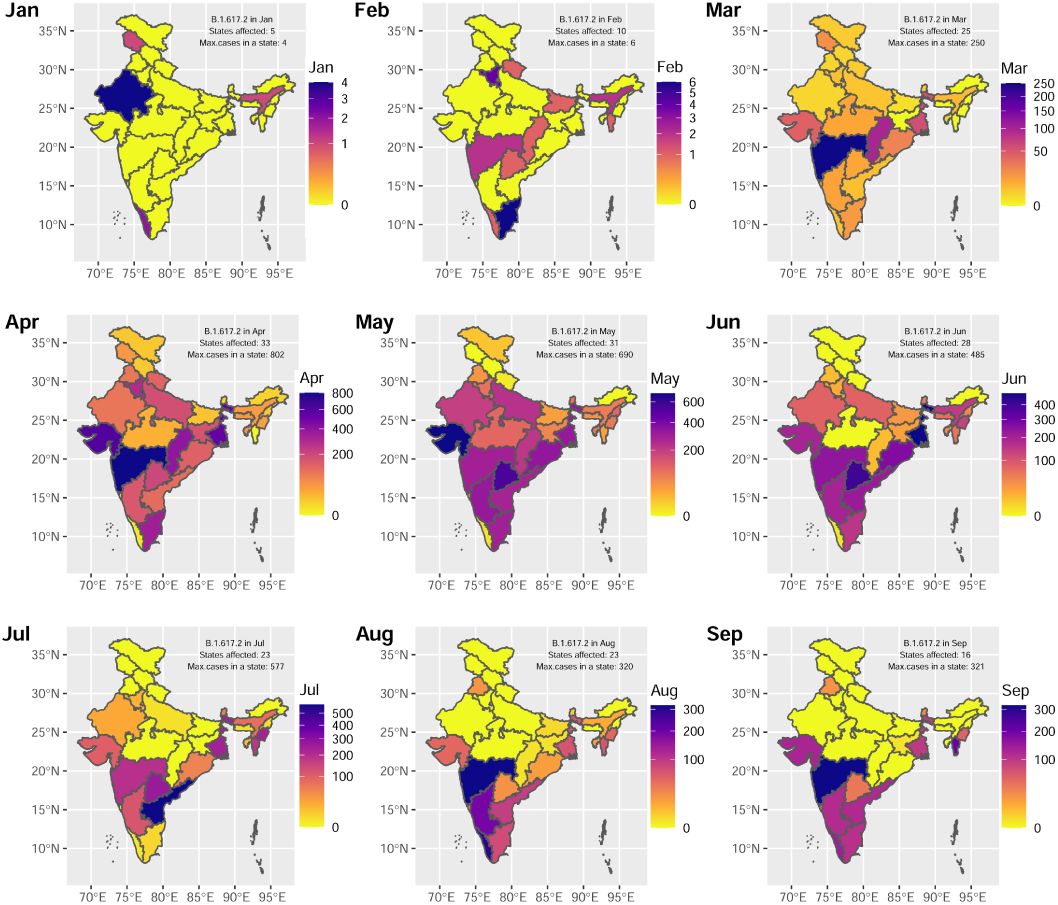
Spatiotemporal transmission/incidences of B.1617.2 (Delta) in Indian states and union territories (UTs). The dynamic incidences of Delta-surge in Indian states and UTs were shown in indicated months). The gradient scale showed the number of events/incidences of Delta for different states and UTs. *Note: The geographical/administrative boundaries are used only for the purpose of data representation*. *The spatial polygonal coordinates or boundaries of state might differ*.

### Correlation of different SARS-CoV-2 lineages and states

A correlation analysis of the VoCs/VoIs was done based on the incidence % per month of different variants of SARS-CoV-2. Here we have studied only the top 10 lineages based on cumulative cases of the variants during January-September 2021. The matrix was plotted on a -1 to +1 scale where positive correlation (>0) and negative correlation (<0) were shown in gradient scale. Zero (0) denoted no correlation. Hierarchical clustering was performed between various VoCs or lineages (Figure 4A). The analysis showed that B.1.617.2 had a high to moderate correlation (0.5-1) based on incidences per month with AY.4, AY.12 and AY.23, indicating the coexistence of Delta and Delta-like. On the other hand, Deltait had negative correlation with other variants (B.1, B.1.1, B.1.1.7, B.1.36, B.1.362.9 and B.1.617.1). Interestingly B.1.617.1 and B.1.17 had high positive correlation among each other and clustered together, but showed moderate correlation with B.1. The third distinguished cluster formed by B.1.36, B.1.36.29, B.1.1 were all positively correlated among themselves, but most distantly related with Delta and Delta-like VoC. The correlation pattern highlighted that Delta and its different sub-lineages (B.1.617.2, AY.4, and AY.23) peak periods/phases is similar and inversely related with all other VoCs/ VoIs based on incidences. In India, delta-surge is the main reason of catastrophic COVID-19 associated deaths. Hence, we studied different states with respective kinetics of Delta-cases associated with the state (Figure 3). In-order to know if there is similar pattern of Delta-surge across the country, we have performed a correlation analysis followed by hierarchical clustering (Figure 4B).

**Fig. 4.**
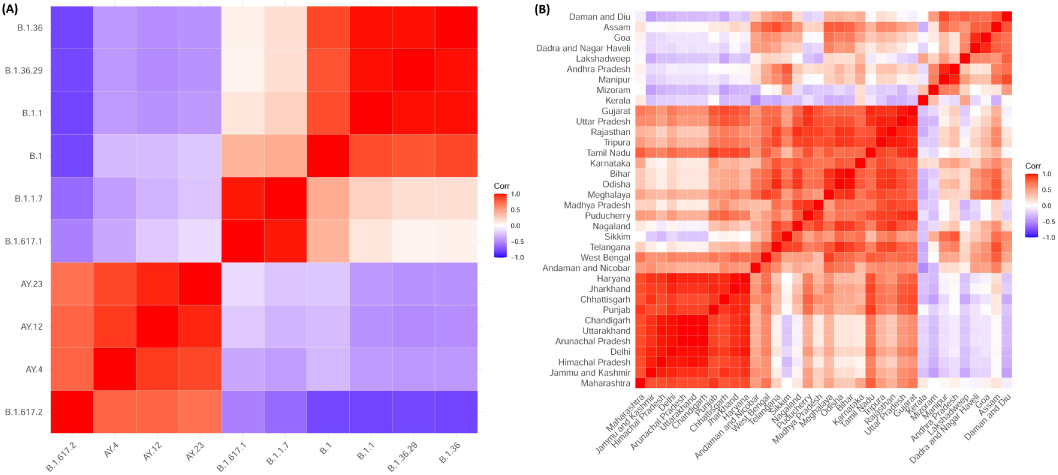
Hierarchical clustering of correlation matrix of various lineages. A; Correlation plot of (A) different variants of SARS-CoV-2 lineages found in Indian population. B; Correlation plot different states/UT with respect to cases of Delta VoC lineage obtained per month

There is moderate to high similarity among the cluster states. Based on K=2, we found 2 clusters as shown in Supplementary Figure S4. The majority of the states fall in cluster 1 whereas cluster 2 formed by states/UT comprising Mizoram, Kerala, Dadra and Nagar Haveli, Goa, Lakshadeep, Manipur, Andhra Pradesh, Assam, Sikkim are distantly related from cluster 1 (Supplementary Figure S4) and possibly less affected by delta-wave (Figure 3).

### Amino acid changes (substitutions/deletions) within the Spike protein of the different variants

In order to study whether the lineages were associated with viral spread (7) and rise in COVID-19 cases, we further consolidate our findings in mutations in the RBD of spike protein (S) (Figure 5A). The spike protein consists of 1273 amino acid of which 319-541 comprises of the RBD region (8). B.1.617.2, AY.4/AY.x, B.1.617.1, B.1, B.1.1.529 (Omicron) and the NC_045512 (wild type/ Reference) denoted as Wt/Ref were selected for the analysis. Fifty genome sequences of each strain having genome length of >29000 were used to obtain consensus nucleotide sequence followed by translated amino acid sequences of the Spike protein. Multiple sequence alignment of all the sequences were done against the reference strain using R package ‘msa’ and the changes/substitutions against each amino acid was identified. Major amino acid changes or substitutions were observed within 19 to 950 region of the S protein. From the analysis it can be observed that B.1 and wt/Ref had similar S protein architecture except the substitution at D614G. All the other variants also showed the change at D614G. Variants B.1.617.1, B.1.617.2 and AY.4/AY.x showed a significant change L452R in RBD site and another at P681R. Interestingly, B.1.617.2 and AY.4/AY.x showed another signature substitution T478K at RBD. Moreover, both the variants (B.1.617/2/AY.x) also had a unique change at D950N. Substitution E156G was unique to AY.4, along with two deletions of amino acids at position 157 and 158 w.r.t. Reference S protein. The mutational analysis further highlighted a completely different architecture of Omicron with 21 signature mutations (unique mutations) as shown by Venn diagram plot (Figure 5A-B). The mutations of Omicron is enriched in the RBD region of the S protein (Figure 5C). In-order to study phylogenetic relationship, pairwise distances from aligned sequences were computed using identity matrix. Neighbor –joining method (9) was applied on these obtained identity matrix. The topology of the tree revealed similarity of S protein origin between NC_04412 and B.1, followed by their close relatedness with B.1.617.1. B.1.617.2 and AY.4 falls under different cluster and are more closely related among themselves. Due to extensive mutations in Omicron Spike protein it falls under separate cluster (Figure 5D).

**Fig. 5.**
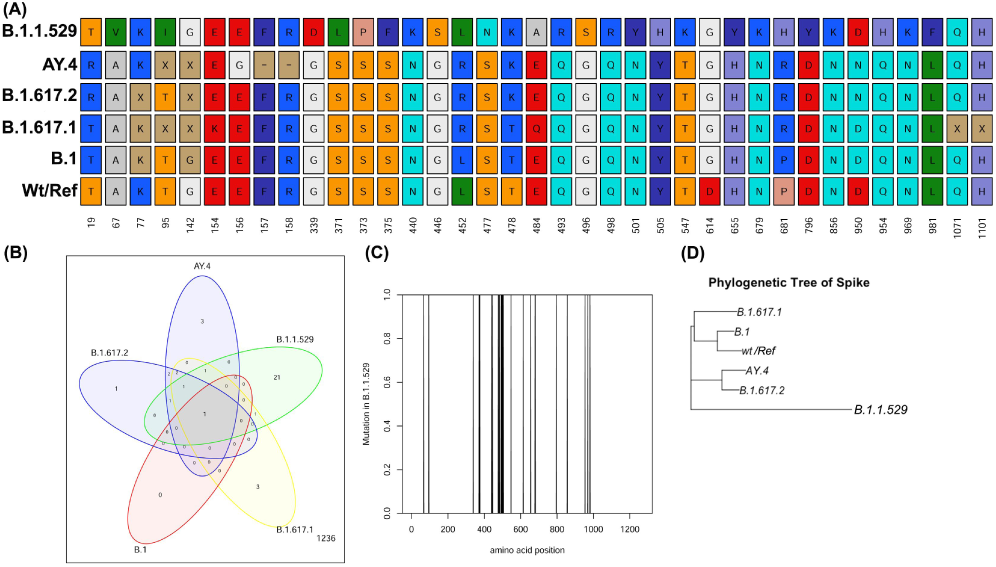
Mutations in various VoC circulating in Indian population in Jan to Sept 2021. A; Mutation in amino acids of various VoCs with respect to wild type/Reference (wt/Ref) S protein is shown along with the position as indicated in number. B; VennDiagram showing overlapping and unique mutations among various VoCs. C; Enriched mutational sites of Omicron (B.1.1.529) lying in the S protein. D; Phylogenetic tree of S protein of various VoCs as indicated.

### Amino acid changes/substitutions in spike (S) protein on SARS-CoV-2 affecting its interaction with ACE-2 receptor and antigenicity

To elucidate the phenotypic effect of amino acid mutations with host cell binding GISAID enabled CoVserver analysis was performed (Table 2) on Delta/Delta-like VoC along with emerging Omicron. Delta/Delta-like variants showed mutations L452R, T478K, D614G, P681R that possibly play role in host-cell interaction as well as and antigenicity. More over entropy was found to be positive (Table 2) indicating that the variant is thermodynamically more stable than wild type strain and might be the possible reason of sudden surge in Delta-Delta like compared to wild type or more closely related VoCs. Interestingly, Omicron, an emerging VoC showed maximum number of amino acid changes in Spike protein enriched in RBD region or its periphery (G339D, S371L, S373P, N440K, G446S, S477N, T478K, E484A, Q493R, G496S, Q498R, N501Y, Y505H, D614G, and P681H) (Table 2), as compared to wild type and Delta and Delta-like. These amino acid positions majorly participate in host cell ACE-2 binding and antibody recognition. Along with this they also help in ligand binding and viral oligomerization (10). The high positive entropy change (Δ=26) for Omicron as compared to reference strain (hCoV-19/Wuhan/WIV04/2019) suggest it to be a more energetically stable and expected to outnumber Delta in near future. The high mutation statistics of Omicron predict that it could be the next VoC potentially responsible for massive surge in COVID cases in India.

**Table 2.** Amino acids involved in phenotypic effects of VoC as compared to Reference.

| VoC | $\Delta S$ | Amino acids for host cell interactions and antigenicity |
| --- | --- | --- |
| B.1.1.529 | 26 | G339D, S371L, S373P, N440K, G446S, S477N, T478K, E484A, Q493R, G496S, Q498R, N501Y, Y505H, D614G, P681H |
| AY.4 | 11 | L452R, T478K, D614G, P681R |
| B.1.617.2 | 8 | L452R, T478K, D614G, P681R |

### Probable new genesis of VoC in near future from Delta or Delta-like

To understand the effect of mutations at the RBD of the Spike protein on the overall interaction, we identified the RDB binding site. Our protein complex search resulted in the structure of SARS-CoV-2 spike receptor-binding domain complexed with ACE2 mutant (PDB ID: 7DMU). First we cross-verified the binding sites using existing protein-protein docking software (11–13). This verification is required since it is known that crystallographic interfaces may not be the biologically functional interface (14). Looking at the effect of the single-point mutation at the RBD stretch of 452-478, we trust on the ProTSPoM web service, which was developed to estimate the effect of single-point mutations on the thermodynamic stability of the spike protein (15). At position 452 and 478 of the receptor binding domain, the sequence alignment indicates existence of mutated amino acids (Figure 5). Our estimation indicates a change in Gibbs free energy while mutating L at 452 positions by R is -0.82 kcal/mol and while mutating T at 478 position by K is -0.12 kcal/mol. Encouraged by this result, at the residue stretch 452-478 we performed a single point mutation by alanine (alanine scanning) of all the residues and tabulated the result in Supplementary Table S4. Apart from 452 and 478, three more positions at the RBD of spike protein may leads to high change in Gibbs free energy viz., Y453, R454, and L461. Interestingly, those three residues occurring within a short span show different chemical property. It might be fascinating to experimentally check the effect of single point mutations at those places with an intention of whether spike protein has possibility of emerging as a variant of concern. In addition, we tested the possibility of emerging single-point deleted variants of the spike protein. We engineer the spike protein by performing single-point deletion within 452-478 residue stretch using an existing method developed by our group (16). However, the study indicates it is on an average with the 50% probability that one single-point deleted variants will assume a folded structure and may emerge as new VoCs or VoIs in future.

## Discussion

The sudden surge of COVID-19 cases in India in 2021 and the devastating second wave corresponds with prevalence of B.1.617.2 (aka Delta variant) and its sub-lineages most significantly AY.4/AY.x (17). Increased incidences of Delta could be linked to its high transmissibility, ability to evade immune responses within human body and diagnostic detection failure (18). High density of population is directly proportional to increased viral replication, mutation and evolution. Such evolutionary processes cause generation of more transmissible and pathogenic viral mutants (19). Dense population of India and poor containment strategies are one of the driving forces behind the deadly second wave. The decline in B.1.617.2 in northern states of India during post 2nd wave may be attributed to herd immunity as those states/ UT are affected mostly during the peak of 2nd wave in India.

Apart from epidemiological considerations the prime cause of Coronavirus transmission and pathogenicity is underlain in its structure. A comprehensive overview of the SARS-CoV-2 structure reveals presence of single stranded positive sense 29.9 kb RNA genome (20). It contains four structural proteins viz. spike (S), envelope (E), membrane (M) and nucleocapsid (N). Apart from structural proteins 16 non-structural proteins (nsp 1-16) help in different viral processing (21). Among all the structural proteins, spike protein is of special research interest because it mediates host cell attachment and entry (21, 22). Detailed structural analysis of homotrimeric S glycoprotein reveals presence of two functional subunits S1 and S2. The S1 contains receptor binding domain (RBD) helping in binding of the virion particle to host cell receptor such as ACE-2 (23). The S2 subunit functions in fusion of virus and host cell membranes (21). Our observations on unprecedented surge of Delta (B.1.617.2) and Delta plus (AY.4/AY.x) in India could be attributed to its unique mutations within the spike protein. The mutation D614G which was observed in all SARS CoV-2 variants was reported to be one of the SNPs that predominated with in all the variants (24). Earlier studies have suggested that the mutation gives a certain replication advantage to the virus. Thus, D614G was associated to increased human transmission and infectivity events (25–27). The spike protein mutation L452R which is found in B.1.617.1, B.1.617.2 and AY.4/AY.x is very important for the pathogenicity of the virus. It confers increased ACE-2 binding affinity and decreased antibody binding capacity, thereby increasing host immune evasion by the virus (28, 29). Mutation P681R have been reported to facilitate S1-S2 cleavage at Furin cleavage site (28). All these mutations facilitate viral replication, transmission and virulence within host cells. Therefore, our findings on predominance of B.1.617.1, B.1.617.2 and AY.4/AY.x over other variants corroborated well with the presence of these mutations within them. Some of the signature mutations of Delta variants are T19R, T478K and D950N (Figure 5), which are present in N terminal domain, RBD and S2 region respectively. Previous literature has highlighted that T19R and T478K (located in epitope binding region) impairs monoclonal antibody mediated neutralization while D950N affects spike protein dynamics thereby increasing virulence (30). So, B.1.617.2 and AY.4/AY.x out-competed B.1.617.1 as well as other variants possibly by evading innate or vaccine induced immunity. In November, 2021 a new variant Omicron (B.1.1.529, BA.1) surfaced with 21 signature and 5 overlapping mutations within its spike protein (Figure 5). The changes in spike protein of omicron can be linked to its increased virulence so surveillance of omicron or omicron-like VoCs will be very important for the coming days. Since our study mainly deals with the genomic surveillance of SARS-CoV-2 during Jan-Sept, 2021 (during onset, peak and post second wave of COVID-19 in India) we have not included Omicron in our surveillance analysis. However, considering the latest scenario of Omicron or Omicron-like VOC surge in several countries including South Africa, UK, USA, etc., tracking of Omicron is needed. Due to its high mutations (30 amino acids changes) within spike protein, the mutational changes of S protein of Omicon-like VoC were analyzed for its role in host-cell receptor binding and antigenicity. There were almost 10-15 (Table 2) amino acids falling in RBD region and its periphery influencing the interactions and antigenicity. Although people in India is immunized against SARS-CoV2 either via mode of vaccination or by herd immunity, the mutations of amino acids might provide an adaptive advantage to overcome immune response or escape antibody neutralization and hence surge in Omicron is anticipated in India too.

Genome surveillance of SARS-CoV-2 associated COVID-19 cases in human population In India showed varying distribution of lineages (VoI/VoC) in 2021 pre, during and post second wave of COVID-19 surge in India. It was evident that variant Delta (B.1.617.2) played major role in the increase in COVID-19 cases and probable reason of second wave in India.The spatiotemporal dynamic transmission/incidences of the Delta or Delta-like VoCs in India were reported for the first time by our group. It indicated that Delta-surge in different states and union territories differ and the north-eastern states and the islands of Indian are less affected by second wave. Considering the recent surge in Omicron-like VoC, and its possible role in altering the host-cell interactions and antigenicity, it’ll be noteworthy to conduct genome surveillance focusing on Omicron-like VoC in parallel with Delta-like VoC. It’ll help in avoiding the stress on the medical systems resulting in fetal consequences due to sudden outburst of incidences of the repetitive waves of COVID-19.

## Data Availability

All data produced in the present study are available upon reasonable request to the corresponding author. S.S

## Acknowledgements

The authors would like to thank CSIR for providing structural and financial support. Sarbar Ali Saha is the recipient of Council of Scientific and Industrial Research (CSIR) Junior Research Fellowship (JRF).

## SUPPLEMENTARY INFORMATION

### Methods

#### Plotting geographical locations/map of the word and India

The spatial polygons or points for world map is obtained using R package (rnaturalearth).The shape file of the state level in India is obtained and downloaded from [GISMAP](https://map.igismap.com/gis-data/india/administrative_boundaries_state_level). The map is solely used for representation purpose of data (COVID-19 related).The plotting was done using R package (ggplot2).The data (table) for COVID-19 cases across the world with days/months/year is downloaded from [WHO COVID-19 Dashboard](https://covid19.who.int/table).

#### Plotting of prevalence of SARS-CoV-2 variant data

All the lineages and their prevalence based on incidences in different states in India in 2021 were ordered with the most prevalent at the top and the least prevalent at the bottom. The top ten lineages based on incidences were selected and scatter plot was constructed using the R/RStudio.

#### Correlation Plot construction and visualization

The correlation matrix based on the prevalence pattern of the lineages was plotted using R/Rstudio. Pearson correlation coefficient method was used for calculating the correlations. Hierarchical clustering of correlation matrix and visualizations of correlation plot was done using R/R studio with package ggplot2.

#### Analysis of amino acid substitutions/changes in the spike protein

The best fifty FASTA nucleotide sequences of genomes of respective SARS CoV-2 variants were downloaded. The selection criteria are: complete genome sequences (>=29000 bases and <1% Ns), high coverage (<1% Ns), exclude low coverage (exclude sequences > 5% Ns) and sequences with entries with complete collection date. The nucleotide sequences were aligned with respect to NCBI Reference Sequence: NC_045512.2 starting at 21563 and extending up to 25384 using R package “‘msa”‘ to obtain a nucleotide consensus sequence representing various lineages. These spike (S) glycoprotein sequence region (21563-25384) was further translated using R package “‘Biostrings”‘. The consensus amino acid sequences of respective lineages were further aligned with amino acid sequences of wild type S protein or NCBI Reference Sequence: YP_009724390.1. The aligned sequences were further analysed for amino acid substitutions/ changes in the different lineages (VoC/VoI) with respect to wild type S protein.

**Table S1.** COVID-19 cumulative cases and death across different countries. The Supplementary data with all the countries can be obtained by sending request to

| Country | Cumulative.cases | Cumulative.deaths | DeathPer10E6Event | income_grp_rank |
| --- | --- | --- | --- | --- |
| Afghanistan | 157951 | 7354 | 46558.74 | 5. Low income |
| India | 34793333 | 479997 | 13795.66 | 4. Lower middle income |
| USA | 51696204 | 808701 | 15643.33 | 1. High income: OECD |

**Table S2.** The metafile of GISAID with aceesion number. The Supplementary data with accession number along with detailed informations can be obtained by sending request to

| Virus.name | Accession.ID | Lineage | Collection.month |
| --- | --- | --- | --- |
| hCoV-19/India/MH-NEERI-NGP-29243/2021 | EPI_ISL_1360324 | B.1.36.8 | 1 |
| hCoV-19/India/WB-1931500693878/2021 | EPI_ISL_1419458 | B.1.1.526 | 1 |
| hCoV-19/India/GJ-GBRC541a/2021 | EPI_ISL_1677770 | B.1.36 | 1 |
| hCoV-19/India/GJ-GBRC541b/2021 | EPI_ISL_1677771 | B.1.36 | 1 |

**Fig. S1.**
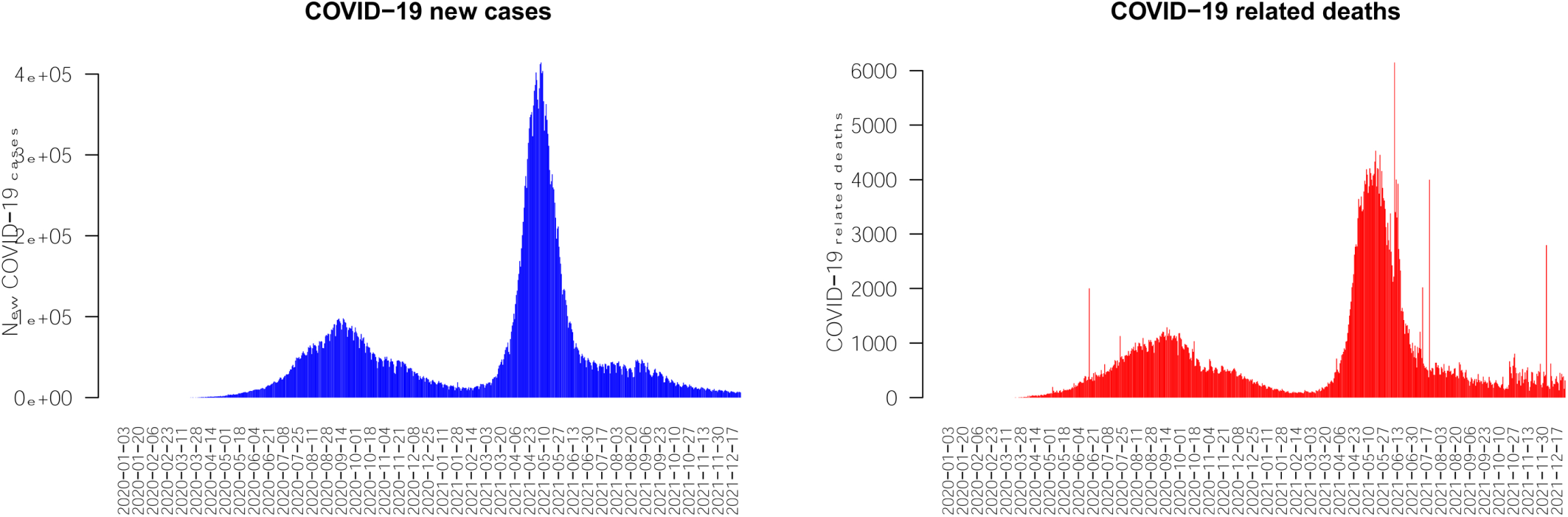
COVID-19 surge/ wave in India.

**Fig. S2.**
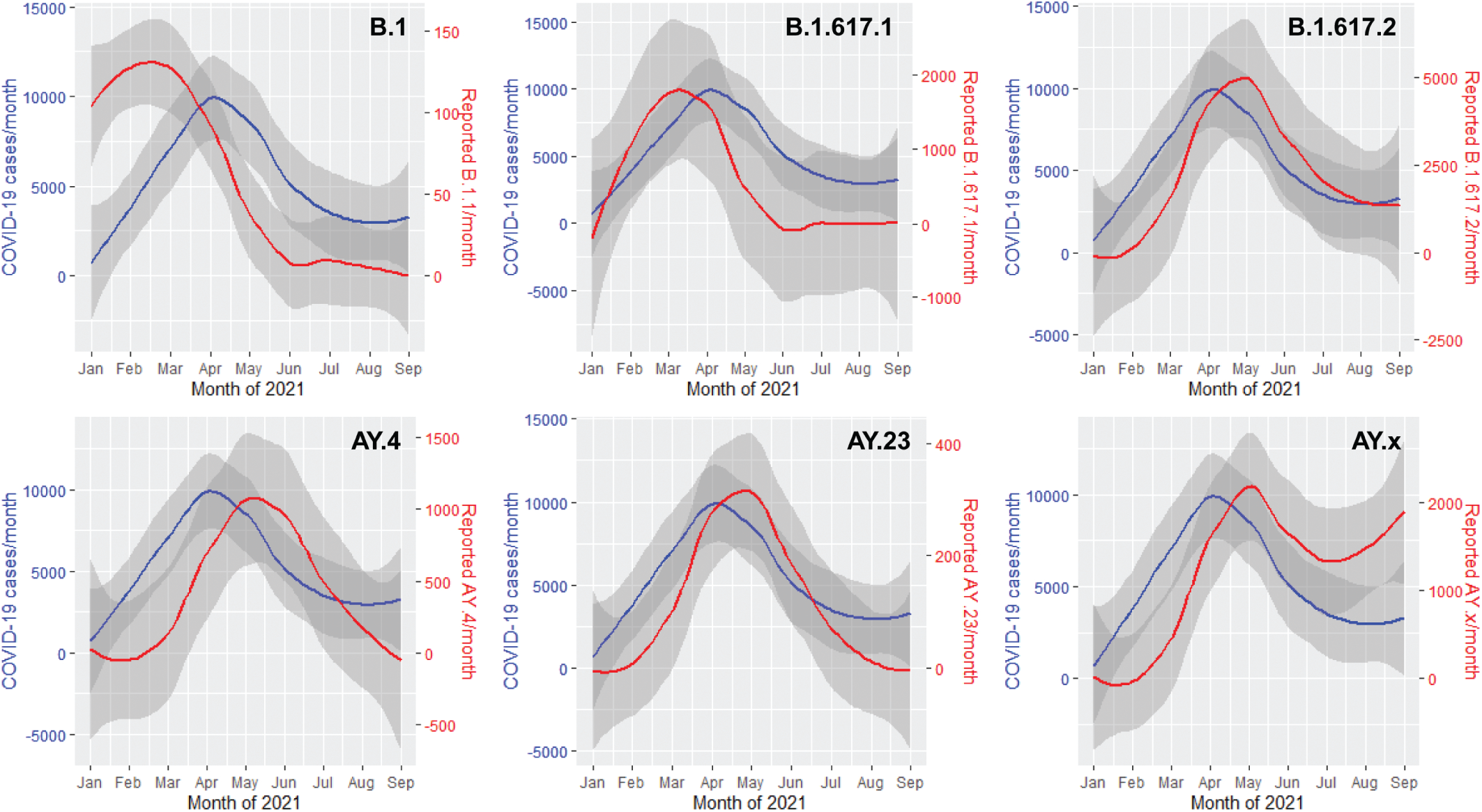
Genome Surveillance of COVID-19 total cases and the respected lineage with respect to months of year 2021. Primary axis (blue colored label) with blue line showing COVID-19 cases observed in the indicated month while secondary y axis (red colored label) with red line indicating reported lineage observed in the indicated month.

**Fig. S3.**
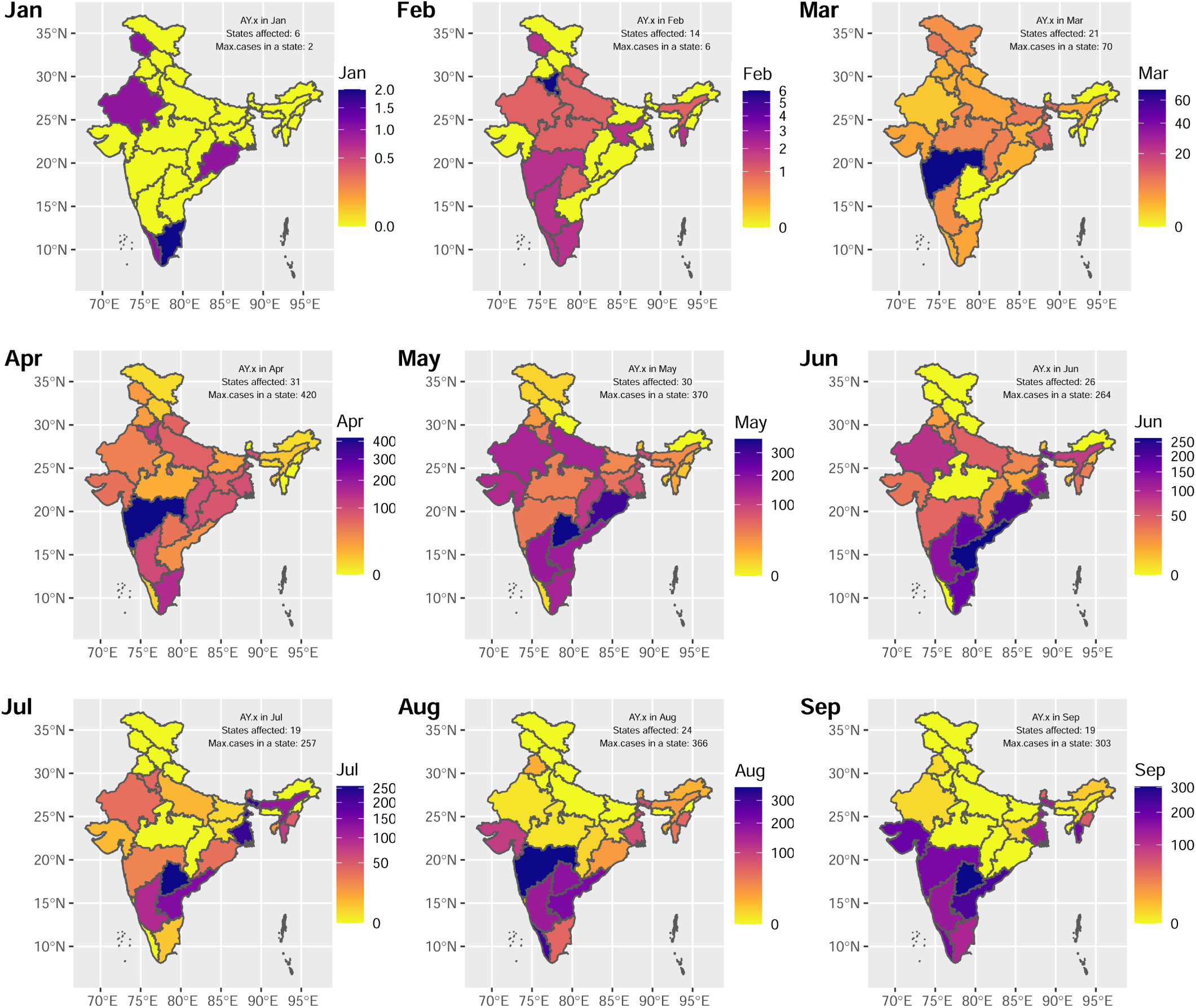
AY.4/AY.x dynamic distribution in Indian states from Jan-September, 2021. The reported cases of AY.x (Delta-like) VoC in respective month in different states of India is plotted. *Note: The geographical/administrative boundaries might differ. The boundaries are used for ease of data representation*.

**Table S3.** The percenatge (%) cases of the lineages from Jan to Sept 2021. The Supplementary data with all lineages circulating in Indian population during Jan to Sept 2021 can be obtained by sending request to

| Lineage | Number | Percentage | Lineage | Number | Percentage |
| --- | --- | --- | --- | --- | --- |
| B.1.617.2 | 18764 | 42.15 | B.1.617.1 | 4478 | 10.06 |
| AY.4 | 3428 | 7.70 | B.1.1.7 | 3213 | 7.22 |
| B.1 | 2640 | 5.93 | AY.12 | 1016 | 2.28 |
| AY.23 | 949 | 2.13 | B.1.36.29 | 660 | 1.48 |
| B.1.1 | 508 | 1.14 | B.1.36 | 488 | 1.10 |
| AY.102 | 440 | 0.99 | AY.43 | 404 | 0.91 |
| AY.16 | 401 | 0.90 | AY.26 | 361 | 0.81 |
| AY.127 | 343 | 0.77 | B.1.1.306 | 293 | 0.66 |
| B.1.1.216 | 278 | 0.62 | AY.20 | 267 | 0.60 |
| B.1.617.3 | 236 | 0.53 | B.1.333 | 211 | 0.47 |
| B.1.525 | 209 | 0.47 | AY.125 | 208 | 0.47 |
| AY.122 | 205 | 0.46 | AY.61 | 197 | 0.44 |
| AY.103 | 177 | 0.40 | B.1.351 | 176 | 0.40 |
| B.1.618 | 164.00 | 0.37 | AY.120 | 159.00 | 0.36 |
| AY.5 | 153.00 | 0.34 | B | 141.00 | 0.32 |
| AY.44 | 132.00 | 0.30 | AY.25 | 130.00 | 0.29 |
| AY.7.1 | 124.00 | 0.28 | B.1.243 | 122.00 | 0.27 |
| B.1.1.526 | 118.00 | 0.27 | AY.50 | 106.00 | 0.24 |
| AY.106 | 104.00 | 0.23 | B.1.36.8 | 100.00 | 0.22 |

**Table S4.** Possible genesis of new VoC due to change in Gibbs free energy (ΔΔG). Position indicates the position of amino acid with respect to wild type Spike protein

| POSITION | RES | $\Delta\Delta G$<br>Kcal/mol | POSITION | RES | $\Delta\Delta G$<br>Kcal/mol | POSITION | RES | $\Delta\Delta G$<br>Kcal/mol |
| --- | --- | --- | --- | --- | --- | --- | --- | --- |
| 452 | L | -1.17 | 453 | Y | -1.87 | 454 | R | -1.95 |
| 455 | L | -1.4 | 456 | F | -1.07 | 457 | R | -1.19 |
| 458 | K | -0.19 | 459 | S | -0.22 | 460 | N | -0.57 |
| 461 | L | -1.61 | 462 | K | -0.19 | 463 | P | -1.28 |
| 464 | F | -1.01 | 465 | E | -0.52 | 466 | R | -0.32 |
| 467 | D | -1.15 | 468 | I | -0.42 | 469 | S | -0.81 |
| 470 | T | -0.07 | 471 | E | 0.06 | 472 | I | -1.43 |
| 473 | Y | -0.84 | 474 | Q | -0.53 | 475 | A | 0 |
| 476 | G | -0.93 | 477 | S | -0.38 | 478 | T | -0.14 |

**Fig. S4.**
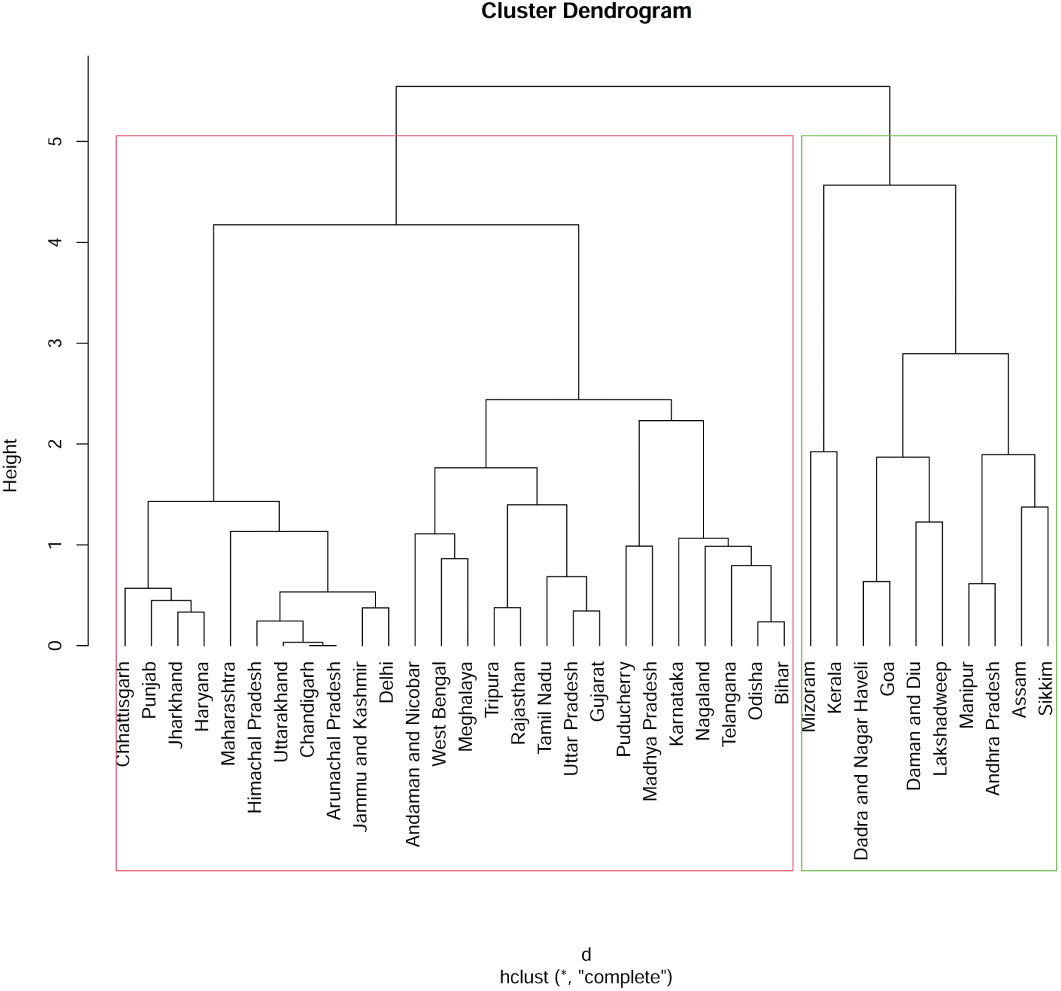
Hierarchical clustering of states /UT based on delta incidences per month.

